# Impact of epidermal melanin on peripheral pulse oximetry measurements

**DOI:** 10.1101/2024.09.17.24313822

**Authors:** James Rice, Jalan Dixon, Ethan Renfrew, Denise Nemeth, Akiva Finkelstein, Yan Gernhofer, Sam Hinckley, Rafael Chaparro, Teresa Samson, Cristina Pizano, Bernard Hildebrand, Steven Mifflin

## Abstract

**Importance:** Pulse oximetry is used widely to assess oxygen saturation in nearly all healthcare institutions.

**Objective:** This study aimed to investigate the relationship between epidermal melanin concentrations, determined by colorimetry, and discrepancies between arterial blood saturation (SaO_2_), determined by arterial blood gas (ABG), and peripheral oxygen saturation (SpO_2_) determined by pulse oximetry.

**Design, Setting, Participants:** This observational, correlational study was conducted over a one-week period in 2024 at the University of the Incarnate Word School of Osteopathic Medicine. Twenty-eight healthy adult volunteers (14 male; 14 female) with varying skin pigmentation were recruited via convenience sampling. Participants with conditions affecting oxygen saturation were excluded.

**Exposure:** Participants underwent non-invasive pulse oximetry to measure peripheral oxygen saturation (SpO_2_) and invasive arterial blood gas (ABG) analysis to measure arterial oxygen saturation (SaO_2_). Melanin concentrations were measured using DSM III ColorMeter on the dorsal and palmar aspects of the hand. SpO_2_ was measured using the Masimo MightyStat^®^ Rx Fingertip Pulse Oximeter, and ABG samples were analyzed using the i-STAT^®^ 1 blood analyzer.

**Main Outcome(s) and Measure(s):** The primary outcome was the difference between SpO_2_ and SaO_2_ across varying melanin concentrations, hypothesized before data collection.

**Results:** Analysis revealed a significant difference between SaO_2_ and SpO_2_ with a moderately positive correlation with average epidermal melanin concentrations (r = 0.432; 95% CI, 0.07 to 0.693; *P* = 0.022) and a moderately inverse correlation with average luminance (r = -0.492; 95% CI,- 0.731 to -0.145; *P* = 0.008). A strong inverse relationship was found between melanin and luminance (r = -0.85; 95% CI -0.928 to -0.698; *P* <0.001). PO_2_ emerged as the leading predictor in two regression models, explaining 50.5% of the variance when combined with melanin.

**Conclusion and Relevance:** Taken together, these data suggest that pulse oximetry overestimates oxygen saturation in individuals with comparatively high epidermal melanin concentrations. Future investigations that include a broader age range and participants with various comorbidities may provide a more comprehensive understanding of the impact of epidermal melanin on the accuracy of pulse oximetry readings.

**Key Points:** *Question:* How does epidermal melanin concentration affect the accuracy of pulse oximetry measurements compared to arterial blood gas results?

*Findings:* This study found that higher epidermal melanin concentrations were associated with greater discrepancies between pulse oximetry and arterial oxygen saturation readings, suggesting that pulse oximeters may overestimate oxygen levels in individuals with darker skin tones.

*Meaning:* Higher epidermal melanin concentrations may result in less accurate pulse oximetry measurements, potentially leading to misinterpretations of oxygen saturation in patients with darker skin tones, raising concerns about the accuracy of these devices across diverse populations.

## INTRODUCTION

Since its introduction in the 1980s, healthcare providers in numerous clinical settings (e.g., hospital wards, outpatient clinics, emergency departments, and prehospital emergency facilities) have relied on pulse oximeters for an accurate determination of a patient’s oxygen saturation. Many important clinical decisions are based on these measurements, and there are many clear benefits to using these devices, including cost-effectiveness, convenience, rapid response time, its non-invasive nature, and its ability to provide continuous monitoring compared to single-point arterial blood gas (ABG) analysis. Although minor discrepancies between measurements of peripheral oxygen saturation (SpO_2_) determined by pulse oximetry and arterial oxygen saturation (SaO_2_) determined by ABG measurements in otherwise healthy individuals has been recognized, inaccuracies reported for patients with one or more comorbidities may have serious consequences. Minor differences in pulse oximetry readings may have a major impact on the decision to provide supplemental oxygen and/or the need for hospitalization, which may have a critical impact on health outcomes and quality of life. The impact of skin color/tone on SpO_2_ measurements determined by pulse oximetry was considered in several earlier studies. These discrepancies depend not only on the specific technology used by pulse oximeters but also on the oxygenation status of the patient. For example, Baek et al.^2^ reported that optical crosstalk found in conventional SpO_2_ sensors led to overestimations of overall oxygen saturation in darker-skinned individuals and that the degree of overestimation was exaggerated in patients with low oxygen saturation. Similarly, reported discrepancies between measurements obtained by two different pulse oximetry devices, notably that the portable devices, although low-cost and widely available, “display worsening agreement with conventional pulse oximeters during hypoxemia.”^3^ Both studies highlight potential flaws in pulse oximeter technology that may delay the appropriate care of patients experiencing hypoxemia in emergent settings.

In 2020, Sjoding et al. published a paper in the *New England Journal of Medicine* entitled “Racial Bias in Pulse Oximetry Measurement” in which they reported that “black patients had nearly three times the frequency of occult hypoxemia as detected by blood gas measurements but not detected by pulse oximetry when compared to white patients.”^3^ This finding prompted a congressional inquiry; in 2021, United States (U.S.) Senators Warren, Wyden, and Booker asked the U.S. Food and Drug Administration (FDA) to conduct a “review of the interaction between a patient’s skin color and the accuracy of pulse oximetry measurements”.^4^ Although they noted that the study was limited given its retrospective design and its reliance on previously collected health data from inpatient hospital stays that could not be statistically correct for all potential founding factors, the FDA emphasized that the findings highlighted a need for further evaluation and an improved understanding of the association between skin pigmentation and the accuracy of pulse oximeter devices.

In response to this evolving narrative, Joe Kiani, the co-inventor of the measure through motion and low perfusion pulse oximeter (Signal Extraction Technology^®^ Pulse Oximeter) and the Founder and Chief Executive Officer of Masimo Corporation also cited the *New England Journal of Medicine* study in response to a changing narrative regarding the ethics of pulse oximetry use in Black patients. According to Kiani, a review of internal data from the Masimo Corporation revealed “a difference between dark and light-skinned subjects of 1%^5^; this value can be compared to a 325% difference reported by Sjoding et al.^3^ However, Kiani mentions in the article that skin color for all subjects included in the validation dataset was recorded using the subjective Melasma Area and Severity Index (MASI) pigmentation scale which scores skin color from 0 to 4 based on perceived visibility of pigmentation. Although Kiani highlighted several potential confounders that might affect the accuracy of pulse oximetry, he failed to mention this subjective scale as a potential source. The use of this type of subjective method to measure skin pigmentation may have a larger effect on the results obtained than those associated with any of the other confounding variables. Moreover, subjective methods of assessing skin pigmentation, including the MASI, can easily introduce variability due to observer bias and environmental factors. The results of studies designed to compared subjective and objective assessments (eg, spectrophotometry) reveal that subjective scales can lack consistency, potentially introducing more error than other confounding variables and may ultimately have a significant impact on the accuracy of important clinical measurements.^6^

A more objective method for categorizing skin color was clearly needed. In addition to the studies discussed above, others who reported that skin color has no significant impact on the accuracy of pulse oximetry typically used the Fitzpatrick or Munsell scales to classify subjects as having a lighter or darker skin complexion. For example, Nickel et al^7^. used the Fitzpatrick scale to show that there was no difference across skin colors in capillary refill time measured by a pulse oximeter when compared to a clinician’s assessment. Another study examined the effects of skin pigmentation on the accuracy of pulse oximetry in infants with hypoxemia using the Munsell system (originally developed to classify the color of soil) to classify skin colors.^8^ Interestingly, Ebmeier et al. found that of the several variables that were significantly associated with differences between SpO_2_ and SaO_2_ readings (eg, body temperature, pulse oximeter model, local factors), the differences were most prominent among that with light as opposed to dark skin color, although “the magnitude of these effects was generally small.”^9^ Instead, of similarly using a categorical variable to measure skin color, this group used the DSM III Skin ColorMeter (Cortex Technology, Hadsund, Denmark) as an objective tool to measure skin color^10^ as a continuous variable and determined the epidermal melanin concentration^11^ in all the study subjects.

## Methods

This study was conducted in accordance with the clinical protocol approved by the Institutional Review Board (IRB) of the University of the Incarnate Word (IRB Reference Number: 2024-1084-FB-v17.6722, Legacy Reference Number: 22-04-004). The study was reviewed under the full board category and received approval on 05/03/2024. The protocol is valid until 03/24/2025.

Findings were collected on 3 separate days over 1 week. Subjects were recruited by word of mouth, email notification and flyers placed in various locations on the University of the Incarnate Word (UIW) campus. Individuals who indicated their interest in participating were emailed an electronic pre-screening questionnaire. Any potential participant who selected yes in response to one or more of the pre-screening questions was excluded. The exclusion criteria included: bleeding disorders (eg, hemophilia, von Willebrand disease, factor V Leiden), dyshemoglobinemias (e.g., anemia, thalassemia, methemoglobinemia, sickle cell disease and/or trait), respiratory pathology (eg, asthma, chronic obstructive pulmonary disease, interstitial lung diseases, cystic fibrosis, emphysema, chronic bronchitis), any skin disorder currently affecting the dorsal or palmar aspects of the hand (e.g., vitiligo, plaque psoriasis, atopic dermatitis, lichenification), or a history of vascular/graft surgery or atrial fibrillation. Also excluded were current smokers (cigarettes and/or e-cigarettes/vapes), subjects with a history of heavy smoking (smoking more than or equal to 25 cigarettes a day), subjects currently taking blood thinners (e.g., aspirin, rivaroxaban, apixaban, clopidogrel, enoxaparin), those with a baseline oxygen saturation below 95%, and subjects without an index that was finger free of nail polish.

The reason for these exclusion criteria was to avoid any possible confounding variables and to limit the risk of ABG-associated complications. Subjects who were not excluded by these criteria and agreed to participate were then allowed to sign up electronically sign up for a time slot on 1 of the 3 available study days. Subjects who participated in the study were given a $10 Starbucks gift card.

Upon arriving at the pre-determined appointment time, the subjects were provided with an informed consent document as approved by the Institutional Review Board (IRB) and were given time to review it, ask questions, and sign the document. The subjects were then escorted into a room and were asked to sit quietly for several minutes before any evaluations were conducted. Topical lidocaine was administered to all subjects while they answered preliminary questions including their name, phone number, age, weight, and height. We used an over-the-counter topical lidocaine formulation (containing 4% lidocaine) applied directly to the skin. The lidocaine was used as a local anesthetic to reduce discomfort during ABG collection. After obtaining each subject’s blood pressure, the Masimo MightyStat^®^ Rx Fingertip Pulse Oximetry unit was then placed on the subject’s left index finger and the SpO_2_ was recorded. This device was selected because it has been described as “the first and only FDA-cleared medical fingertip pulse oximeter available Over-The-Counter (OTC) direct to consumers without a prescription”, contains “technology relied on by hospitals and clinics around the world to monitor more than 200 million patients every year”, and was “shown to have no clinically significant difference in accuracy or bias between light- and dark-skinned individuals.”^12^ A DSM III Colorimeter was then placed on the dorsal aspect of the left hand and data on melanin concentration, erythema, luminance, red/green chroma, and yellow/blue chroma were collected. These measurements were then repeated on the palmar aspect of the left hand as well. The Colorimeter allows for objective quantification of participant skin pigmentation. According to a recent publication by Okunlola et al. that proposed changes to current pulse oximetry research protocols, “By objectively quantifying skin color, researchers can better distribute study subject representation to accurately account for the spectrum of skin pigmentation.”^13^ To facilitate a well-controlled testing environment, the subject was then immediately escorted to another room for ABG analysis. This was done in accordance with Okunlola et al. who noted that, “SaO2 is prone to fluctuations over short periods of time, necessitating that the blood sampling and the pulse oximeter reading be done as close together as possible for accurate comparisons.”^13^ An Allen test was performed to assess for collateral circulation and capillary refill time was noted. Ultrasound guidance was available to assist in locating the radial artery if needed. Any arterial blood sample that was contaminated was immediately discarded. Each subject was allotted a maximum of two attempts for arterial blood draw. The area was sterilized using a standard disinfectant wipe commonly used for arterial blood gas (ABG) draws. The procedure was performed by two medical students under direct supervision from chief internal medicine residents and the faculty clinical sponsor, a board-certified rheumatologist. To ensure hemostasis, subjects were instructed to remain still and apply pressure to the puncture site for 5 minutes following the procedure. Each subject was contacted the next day to confirm there were no complications, and none were reported. Immediately after a successful arterial blood draw was obtained, the sample was analyzed with the i-STAT® 1 blood analyzer and CG8+ cartridges, and the following values were obtained: pH, the partial pressure of carbon dioxide, CO_2_ content, sodium, potassium, calcium, glucose, hemoglobin, hematocrit, base excess, bicarbonate, SaO_2_, and partial pressure of oxygen (pO_2_).

## RESULTS

Thirty-nine subjects volunteered for the study; 11 were disqualified for various reasons, including: wearing nail polish (n = 2), the inability to draw arterial blood after two attempts (n = 8), and the blood sample clotting quickly, preventing analysis on the i-STAT 1^®^ device (n = 1). Data was collected successfully from 28 participants (14 males and 14 females) with an average of 35.5 +/-10.2 years. All participants were students or faculty of the UIW School of Osteopathic Medicine. Data from these 28 individuals were used to determine the difference between SpO_2_ measurements obtained by pulse oximetry and SaO_2_ determined by ABG and examined the change in SaO_2_ vs SpO_2_ in relation to the measured physical characteristics of the skin using Pearson correlation (**Table 1**). The results revealed a moderate positive correlation with average melanin (r = 0.432; 95% CI 0.07 to 0.693; *P* = 0.022) and average erythema (r = 0.394; 95% CI 0.024 to 0.669; *P* = 0.038), and a moderate inverse correlation with average luminance (r = -0.492; 95% CI -0.731 to -0.145; *P* = 0.008). Average luminance was also inversely correlated with SaO_2_ vs SpO_2_ change (r = -0.492; 95% -0.731 to -0.145; *P* = 0.008) and average melanin (r = -0.85; 95% CI -0.928 to -0.698; *P* < 0.001). Additional data collected included subject age, body mass index (BMI), and PO2; only the correlation between PO2 values and SaO_2_ vs SpO_2_ changed reached statistical significance (r = 0.616; 95% CI -0.804 to -0.316; *P* < 0.001).

**Table 1.**
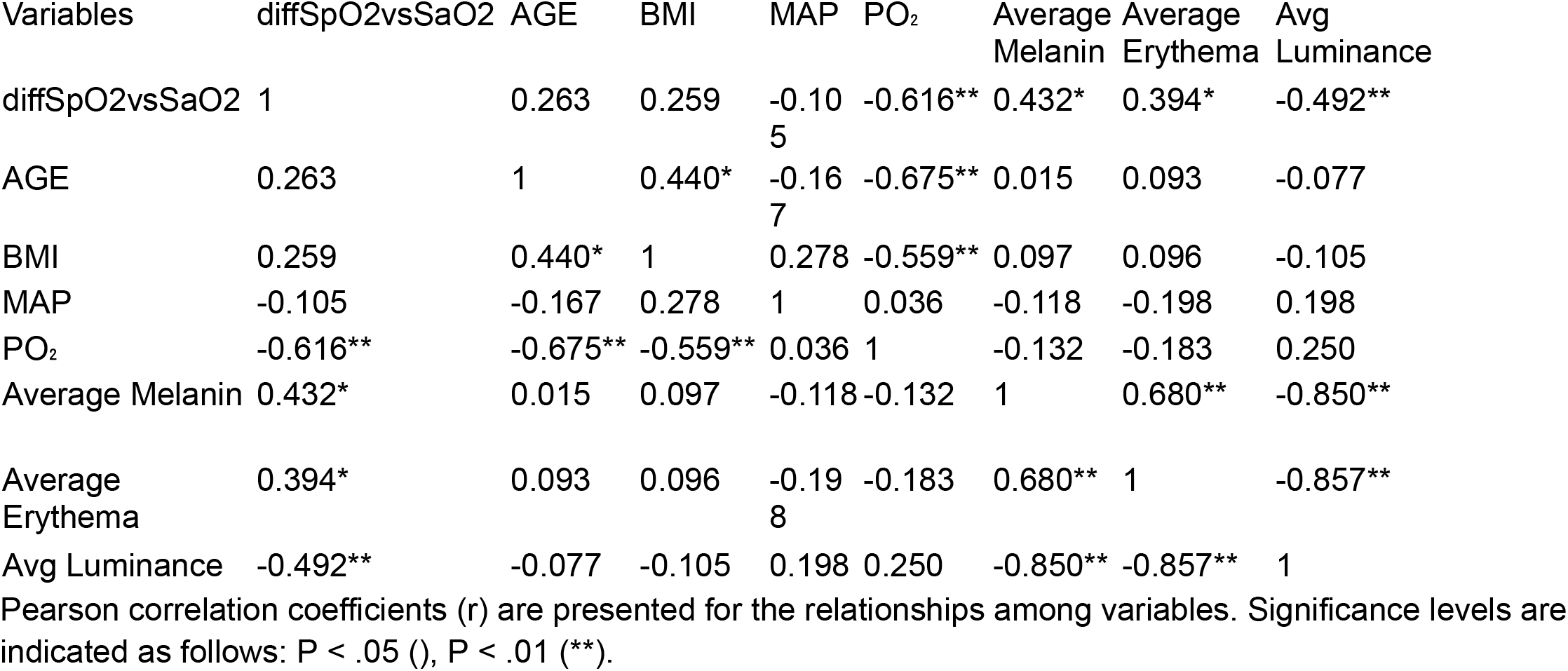
Pearson Correlations of Key Variables.

These interactions were explored further using a stepwise logistic regression with change in SaO_2_ vs SpO_2_ as the dependent variable. Two independent models were developed: Model 1 (PO_2_) and Model 2 (PO_2_ + average melanin). Model 2 provided a better fit, as it explained 50.5% of the data variability (R^2^ = 0.505, Akaike information criterion (AIC) = -7.700, *P* = 0.019) compared to 38% in Model 1 (R^2^ = 0.380, AIC = -3.386, *P* < 0.001), as shown in **Table 2**.

**Table 2.**
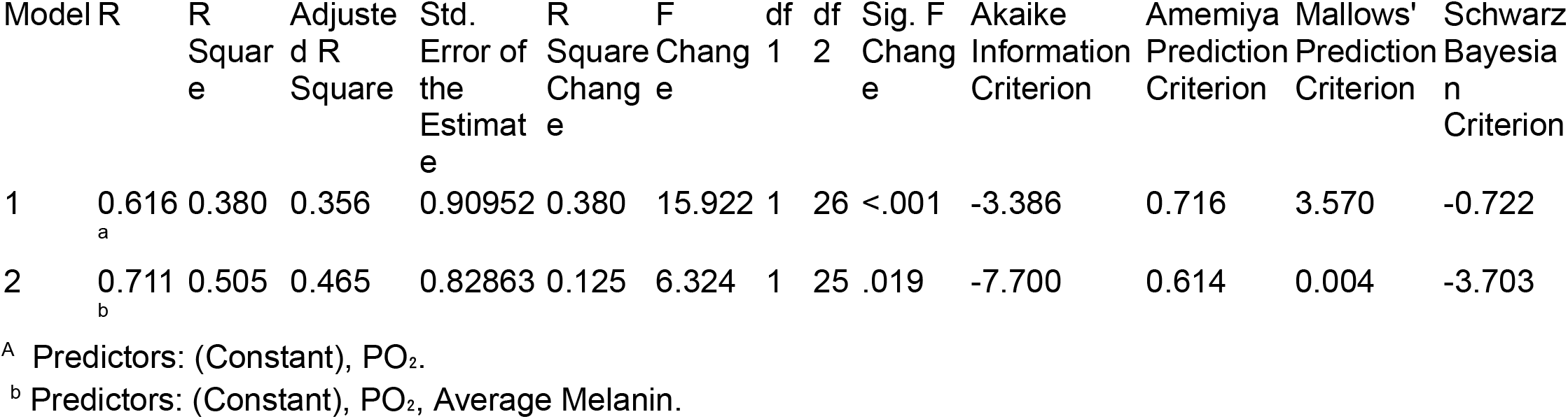
Model Summary for Predictors of Stepwise Regression.

Collectively, the results reveal a significant discrepancy between measurements of pO_2_ in the blood as determined by pulse oximetry (SpO_2_) when compared to ABG (SaO_2_).

## DISCUSSION

The findings of this study provide critical insights into the relationship between epidermal melanin concentration and the differences in oxygen saturation readings, suggesting that this may be a factor contributing to the significant discrepancies between peripheral oxygen saturation (SpO_2_) as measured by pulse oximeters and arterial blood hemoglobin saturation (SaO_2_) determined by arterial blood gas (ABG) analysis. The results demonstrate that higher melanin concentrations are associated with larger disparities between SpO_2_ and SaO_2_ readings and suggest that pulse oximetry may overestimate oxygen saturation in individuals with darker skin tones. This observation aligns with previous research, which has pointed to inaccuracies in pulse oximetry readings for individuals with higher levels of skin pigmentation, potentially contributing to racial disparities in clinical decision-making.

Our correlation analysis showed a moderately positive relationship between average melanin concentration and the discrepancy between SaO_2_ and SpO_2_. There was a moderate inverse correlation between average melanin concentration and luminance, a measure inversely related to skin pigmentation. These findings are consistent with the hypothesis that high concentrations of epidermal melanin can interfere with the accurate measurement of oxygen saturation by pulse oximeters.

Stepwise regression analysis of the data further highlighted the importance of considering both PO_2_ and melanin concentration when predicting a potential discrepancy between SaO_2_ and SpO_2_. The inclusion of melanin as a predictor significantly improved the model’s explanatory power, indicating that skin pigmentation contributes critically to the variability of pulse oximetry readings. This finding has important clinical implications, as it suggests that pulse oximeters may not provide readings that are equally reliable readings across different populations and that current technology may lead to misdiagnosis or delayed treatment in individuals with darker skin tones.

These findings have several implications for clinical practice and the design of pulse oximetry devices. First, healthcare providers need to be aware that pulse oximetry readings may be inaccurate in patients with higher epidermal melanin concentrations. In cases in which precise oxygen saturation measurements are critical, it may not be sufficient to rely solely on pulse oximetry. ABG analysis should be considered as a confirmatory tool, particularly in patients with darker skin tones

Second, these results underscore the necessity for further research and development in the field of pulse oximetry. Manufacturers should consider refining the algorithms and sensor technologies currently used in pulse oximeters to minimize the impact of skin pigmentation on measurement accuracy. This may involve the development of devices that are calibrated to account for a wider range of skin tones, thereby reducing the inaccuracies currently observed in SpO_2_ readings in patients with darker skin.

Finally, this study emphasizes the importance of using objective and precise methods for categorizing skin color in research. In contrast to previous studies that relied on subjective scales such as the Fitzpatrick scale or the Munsell system, the use of a colorimeter to measure epidermal melanin concentration provided a more accurate and reliable assessment of skin pigmentation. This approach not only enhances the validity of the findings, but also sets a standard for future research in this area.

This study does include some limitations. The sample size was relatively small with a group of participants of similar age and health status, and the study was conducted at a single location. This may limit the generalizability of the findings to broader and more diverse populations. Additionally, while this study focused on the relationship between epidermal melanin concentration and the accuracy of pulse oximetry, other factors such as environmental conditions, device-specific variables, and patient movement during measurement were not extensively explored and could also contribute to the discrepancies observed.

## CONCLUSION

In conclusion, this study provides compelling evidence that pulse oximetry may overestimate oxygen saturation in individuals with higher melanin concentrations, raising concerns about the reliability of these devices across different skin tones. These findings raise important concerns about the reliability of these devices as a basis for the clinical evaluation of diverse populations. These results emphasize the need for increased awareness among healthcare providers, further research into the development of more accurate pulse oximetry technologies, and the adoption of objection methods for assessing skin pigmentation in clinical and research settings.Furthermore, these findings highlight the need for additional research involving subjects from a wide range of age groups and a broader spectrum of comorbidities. A follow-up study is planned that will be designed to improve our understanding of the clinical implications of the observed discrepancies in oxygen saturation measurements and to explore additional variables that may affect the accuracy of pulse oximetry, particularly in subjects with concurrent comorbidities.

As healthcare continues to evolve towards personalized medicine, it will be critical to ensure that diagnostic tools, including pulse oximeters, provide accurate readings across all members of diverse populations. This will improve clinical outcomes, reduce unnecessary medical expenses, and alleviate the financial burden on the healthcare system by reducing the incidence of misdiagnosis and inappropriate treatments. By continuing to explore and address the limitations identified in this study, the medical community can take meaningful steps towards more equitable and cost-effective healthcare for all patients.

## Data Availability

All data produced in the present study are available upon reasonable request to the authors

